# First evidence of microplastics in human ovarian follicular fluid: an emerging threat to female fertility

**DOI:** 10.1101/2024.04.04.24305264

**Authors:** Luigi Montano, Salvatore Raimondo, Marina Piscopo, Maria Ricciardi, Antonino Guglielmino, Sandrine Chamayou, Raffaella Gentile, Mariacira Gentile, Paola Rapisarda, Gea Oliveri Conti, Margherita Ferrante, Oriana Motta

**Author notes:** Correspondence to: L. Montano, Andrology Unit and Service of Lifestyle Medicine in UroAndrology, Local Health Authority (ASL) Salerno, Coordination Unit of the Network for Environmental and Reproductive Health (Eco-Food Fertility Project), “S. Francesco di Assisi Hospital”, 84020 Oliveto Citra, SA, Italy. E-mail addresses (L. Montano) and. These authors contributed equally as co-first (Luigi Montano, Salvatore Raimondo). These authors contributed equally as co-last (Margherita Ferrante, Oriana Motta).

## Abstract

Plastic pollution is a pressing global issue, with over 400 million tons produced annually and projections of 1.1 billion tons by 2050. Microplastics (MPs), ranging from 5 mm to 1 µm, are pervasive in the environment. They are found in air, sea, freshwater, soils, food chains and studies show that tiny MPs, smaller than 10 μm, can cross cellular membranes, posing potential health risks through oxidative stress, inflammation, immune dysfunction, neurotoxicity and reprotoxicity.

In recent years, research has shown that microplastics have negative effects on the female reproductive systems of animals. However, there is still a lack of evidence on how the accumulation of microplastics affects the reproductive health of human females. This study aimed to examine the presence of microplastics in the ovarian follicular fluid of 18 women undergoing assisted reproductive treatment whose samples were processed using a patented method endorsed nationally and internationally. Plastic particles <10 µm were measured using SEM with EDX detection. Preventive measures were taken to avoid contamination during the process. Microplastics (dimensions <10 µm) were detected in 14 out of 18 samples of follicular fluid, with an average of 2191 p/ml (0 - 7181p/ml) and with a mean diameter of MPs of 4.48 µm (3.18-5.54 µm). A significant correlation was found between microplastic concentration and FSH (p-value <0.05), as well as a weak correlation with BMI, age and Estradiol. There was no correlation with fertilization outcomes, miscarriages, or live birth. This is the first study to provide evidence for microplastics’ presence in ovarian follicular fluid in women undergoing assisted reproductive treatment, representing a potential threat to female reproductive function.

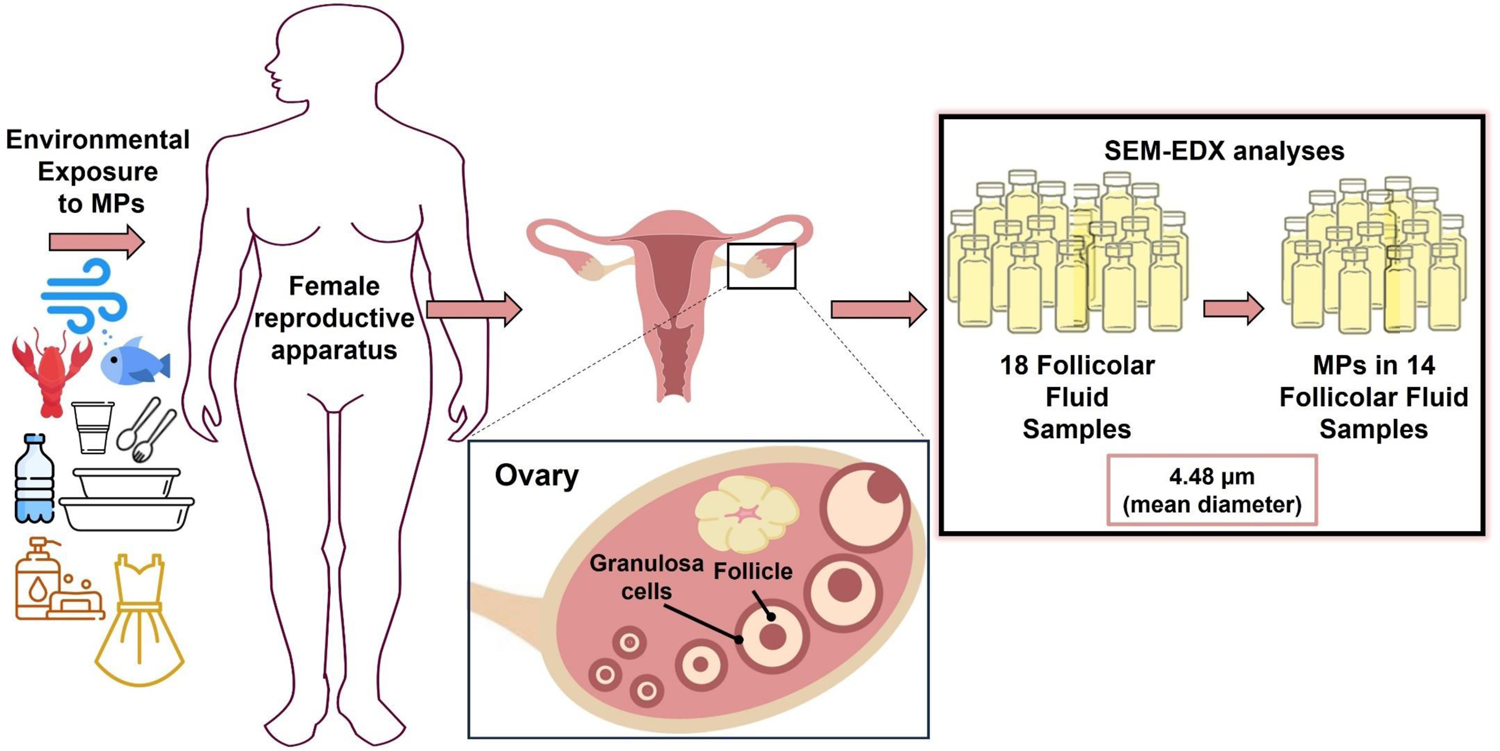

## 1. Introduction

The increasing presence of plastic and its degradation products in the environment has become a global emergency. Every year, over 400 million tons of plastic are produced and this number is expected to increase even further, being highly likely that by 2050, annual production will reach 1.1 billion tons (UNEP, 2021). The pervasiveness of plastic is deeply influencing the planet’s ecosystem and biogeochemical cycles to a point where it has become a ubiquitous and distinctive element in the Earth’s geology at every latitude, so much so that the term “Plasticene” can be defined as the geological era characterized by this massive presence (Rangel-Buitrago and Neal, 2023).

From the degradation process of plastic, particles of plastic on a micrometric scale are created, known as microplastics, ranging from 5 mm to 1 µm in size, and nanoplastics with a diameter smaller than 1µm. Microplastics (MPs) can have different shapes (fibers, fragments, spheres, beads, films, scales, pellets, and foam) and colors depending on the original form of the large plastics from which they derive. MPs derived by fragmentation of plastic wastes are called “secondary” MPs, while “primary” MPs are specifically produced by humans both for their abrasive properties and to improve the stability of certain products, consequently MPs are intentionally added in cosmetics, toothpaste, household products, detergents, and paints formula (Ricciardi et al., 2021). Due to their small size, they are transported everywhere, and therefore we find them increasingly in the air, sea and freshwaters, soils, and being bioconcentrated in the food chain (Ferrante et al., 2022; Garrido Gamarro, 2022; Liu et al., 2023; Oliveri Conti et al., 2020) MPs reach humans (Pironti et al., 2021).

There is a growing concern regarding human health. MPs primarily enter the human body through the ingestion of food and beverages, as well as through inhalation, and finally through skin absorption of MPs contained in cosmetics (Yang et al., 2023). Especially, the large consumption of water mineral PET bottles results in a concentration of MPs between 110,000 and 370,000 particles (90% are nanoplastics and 10% microplastics) (Qian et al., 2024). Thanks to the use of an extremely sensitive methodology, the Estimated Daily Intake (EDI) of 1,531,524 p/kg/body-weight/day, corresponding to 40.1 μg/kg/body-weight/day, and 3,350,208 p/kg/body-weight/day, corresponding to 87.8 μg/kg/body-weight/day, was evaluated. Several studies on mammals indicate that MPs smaller than 10 μm can cross cellular membranes, posing potential health risks through oxidative stress, inflammation (Pulvirenti et al., 2022), immune dysfunction (Yang et al., 2022), neurotoxicity (Wang et al., 2022), altered biochemical and energy metabolism, impaired cell proliferation, gut microbiota alteration (Huang et al., 2021) or disrupted microbial metabolic pathways, abnormal tissue development, and carcinogenicity (Haddadi et al., 2022; Najahi et al., 2022). Evidence continues to accumulate in this regard, particularly concerning endocrine disruption (Ullah et al., 2023), and reproductive toxicity (Huang et al., 2023).

Furthermore, MPs, due to their hydrophobic surface, act as a Trojan horse for other types of notoriously toxic environmental contaminants (such as dioxins, polychlorinated biphenyl ethers, bisphenols, phthalates, polybrominated diphenyls, polycyclic aromatic hydrocarbons, and heavy metals) which can bind to them and, through processes of bioaccumulation and biomagnification, cause additional harm to living organisms through synergistic effects (Schell et al., 2022; Ullah et al., 2023). Beyond that, nano and MPs can serve as vehicles for microorganisms and promote infections (Beans, 2023).

All of this toxic burden to which the population is increasingly exposed represents one of the reasons why in recent decades there has been a decline in human fertility (Aitken, 2022; Pan et al., 2024). A recent report from the World Health Organization (WHO) estimates a global prevalence of 17.5% of couple infertility (WHO, 2023) and a recent meta-analysis has recorded a global decrease in total sperm count of 62.3% from 1973 to 2018 (Levine et al., 2023). Moreover, this has also been accompanied by an increase in the incidence of testicular tumors and a decrease in testosterone, because the male reproductive system is particularly sensitive to environmental contaminants (Gallo et al., 2020; Montano, 2020; Montano et al., 2018). However, there are areas within the same country or region that have higher pollution rates, accompanied by a higher incidence of reproductive problems for both males (Bergamo et al., 2016; Ferrero et al., 2024; Lettieri et al., 2020) and females (Ding et al., 2022; Xue et al., 2021).

Therefore, the significant impacts that environmental contaminants have on reproductive function are alerting the scientific community and policymakers to the increasing burden and potentially adverse effects on human health, starting from the reproductive health of emerging contaminants such as nano and MPs. In this regard, the discovery of MPs in the human seminal fluid has greatly heightened these concerns (Montano et al., 2023). Although there is currently a lack of evidence of reproductive effects in humans, several studies on animals demonstrate significant alterations in male reproductive function. In male mice exposed to polystyrene particles (PS-MPs 5.0-5.9 μm) in saline solution for six weeks, a reduction in sperm motility, an increase in abnormal sperm forms, and a decrease in testosterone levels were observed (Coffin et al., 2022).

Another study following exposure to PS-MPs observed processes of desquamation, atrophy, and apoptosis of germ cells in much of the seminiferous epithelium with increased levels of interleukins, effects closely related to the nuclear factor erythroid 2-related factor 2 (Nrf2)/heme oxygenase-1 (HO-1)/nuclear factor-kappa B (NF-κB) signaling pathway (Hou et al., 2021). As revealed in an in vivo study on mouse sperm, polystyrene nanoplastics (PS-NP) trigger elevated ubiquitination of Ras-related C3 botulinum toxin substrate 1 (RAC1) and cell division cycle 42 (CDC42) (Xu et al., 2023). The calculated minimal human equivalent MPs dose causing a reduced semen quality was 0.016 mg/kg/day (Zhang et al., 2022).

On the female sex, ovarian functionality is also particularly sensitive to the effects of various endocrine disruptors that induce reproductive health issues, such as infertility, imbalances in sex hormones, and premature ovarian insufficiency (Ding et al., 2022). Although there is a lack of studies on the effects of MPs on humans, there is evidence from animal models demonstrating their adverse effects. In a study on mice, after administration of polyethylene MPs (PE-MPs) of 10-150 μm, (40 mg/kg/day) for 30 days, reduced oocyte maturation was observed, with a decreased capacity for fertilization of these oocytes, alterations in the development of the resulting embryos, related to oxidative stress damage with implications for DNA and mitochondrial dysfunction in the exposed oocytes (Zhang et al., 2023). Another study has shown a reduction in plasma levels of 17β-estradiol (E2) and testosterone (T) in female *Oryzias melastigma* after 60 days of exposure to PS-MPs (Wang et al., 2019). Exposure to 0.5 μm PS-MPs (0, 0.015, 0.15 and 1.5 mg/day for 90 days) has been shown to induce overproduction of reactive oxygen species (ROS) and thus oxidative stress contributes to ovarian tissue alterations (An et al., 2021).

In any case, these findings in mammals lead us to consider that nano and microplastics can also accumulate in ovarian tissue and therefore produce adverse effects on female fertility.

The main purpose of this preliminary study was therefore to verify the presence of microplastics in the follicular fluid of 18 women undergoing assisted reproduction according to a protocol developed by some of the authors. This study, to the best of our knowledge, represents the first evidence of MPs in human follicular fluid. Although still limited in numbers, this discovery should serve as an important warning signal about the invasiveness of these emerging contaminants in the female reproductive system, considering that they can alter its composition and have an impact on the oocyte (Gosden et al., 1988; Petro et al., 2012), thus posing a significant reproductive risk for our species.

## 2. Materials and methods

### 2.1 Patients’ enrolment

The study was performed in accordance with the guidelines and regulations described by the Code of Ethics of the World Medical Association (Declaration of Helsinki) and falls within the scope of the EcoFoodFertility project (www.ecofoodfertility.it, accessed on 08 Febr 2024), approved by the Ethical Committee of the Local Health Authority Campania Sud-Salerno (Committee code n. 43 of 30 June 2015). EcoFoodFertlity is a human biomonitoring project which is investigating the presence of various contaminants in biological fluids and their potential effects on reproductive health. All patients were fully informed about the project and signed an informed consent to participate. N.18 ovarian follicular samples were collected from women undergoing assisted reproductive treatment at IVF (In Vitro Fertilisation) center of Mediterraneo PMA (medically assisted procreation) in Salerno (Campania Region, Southern Italy) between February 2019 and January 2020. Every participant had declared to consume plastic packaged food and plastic bottled water or beverages during the three years before sampling. N.4 women out of 18 participants had secondary infertility. The primary infertility causes included declined ovarian reserve (DOR), polycystic ovary syndrome (PCOS), tubal factors, advanced age, and unexplained infertility.

The principal characteristics of study participants can be summarized in Table 1. More detailed information on all participant parameters can be found in the supporting material.

**Table 1.**
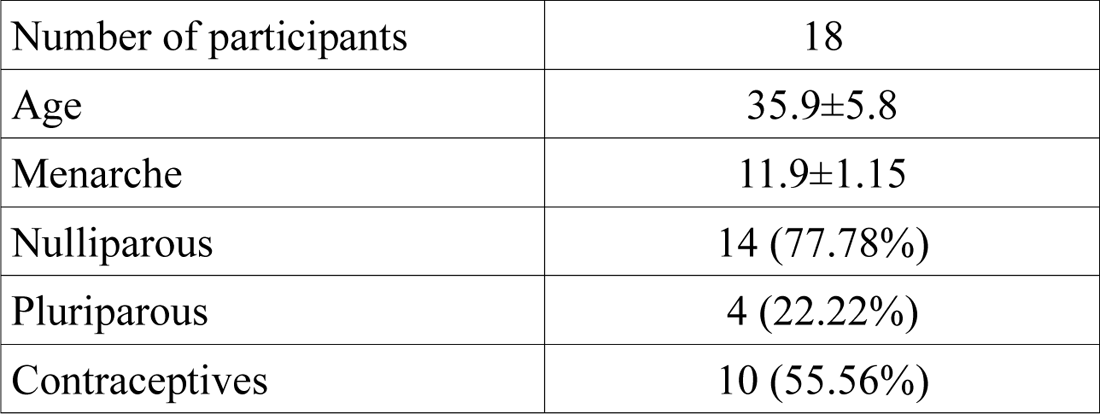
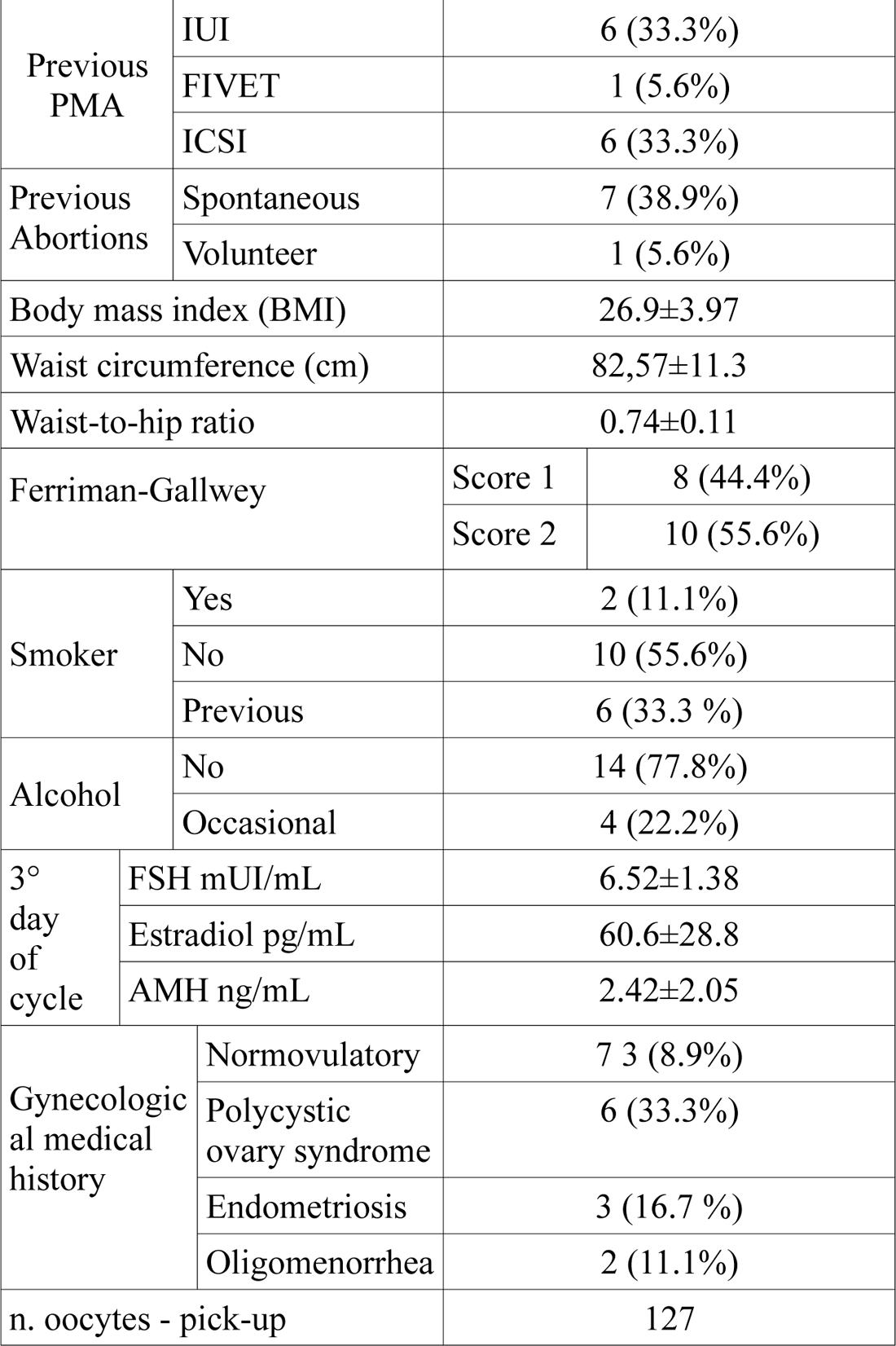
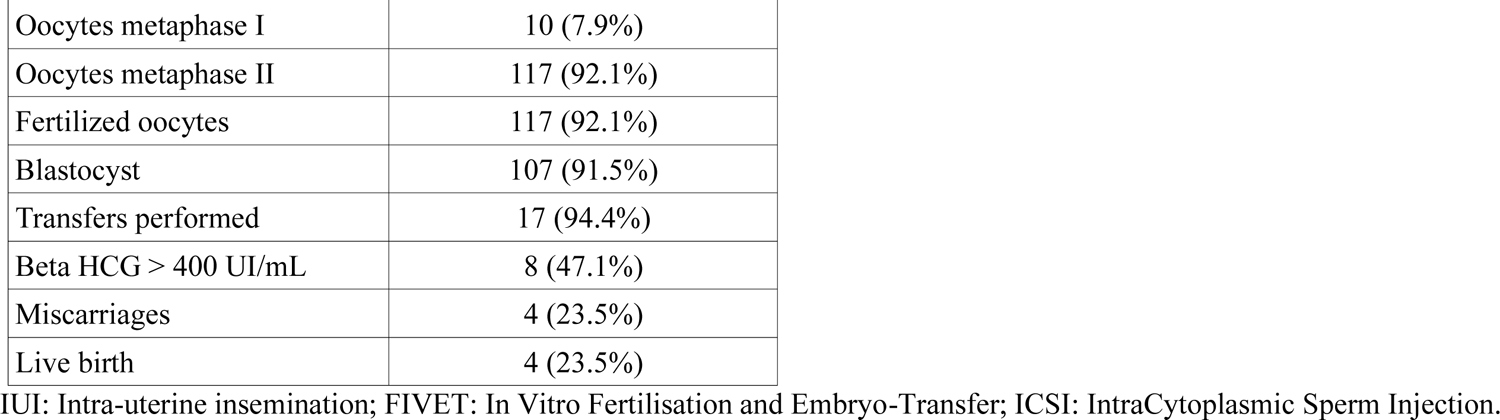
Main characteristics of study participants.

|  |  |
| --- | --- |
| Number of participants | 18 |
| Age | 35.9±5.8 |
| Menarche | 11.9±1.15 |
| Nulliparous | 14 (77.78%) |
| Pluriparous | 4 (22.22%) |
| Contraceptives | 10 (55.56%) |

|  |  |  |  |
| --- | --- | --- | --- |
| Previous PMA | IUI | 6 (33.3%) |  |
|  | FIVET | 1 (5.6%) |  |
|  | ICSI | 6 (33.3%) |  |
| Previous Abortions | Spontaneous | 7 (38.9%) |  |
|  | Volunteer | 1 (5.6%) |  |
| Body mass index (BMI) |  | 26.9±3.97 |  |
| Waist circumference (cm) |  | 82,57±11.3 |  |
| Waist-to-hip ratio |  | 0.74±0.11 |  |
| Ferriman-Gallwey |  | Score 1 | 8 (44.4%) |
|  |  | Score 2 | 10 (55.6%) |
| Smoker | Yes | 2 (11.1%) |  |
|  | No | 10 (55.6%) |  |
|  | Previous | 6 (33.3 %) |  |
| Alcohol | No | 14 (77.8%) |  |
|  | Occasional | 4 (22.2%) |  |
| 3° day of cycle | FSH mUI/mL | 6.52±1.38 |  |
|  | Estradiol pg/mL | 60.6±28.8 |  |
|  | AMH ng/mL | 2.42±2.05 |  |
| Gynecologic al medical history | Normovulatory | 7 3 (8.9%) |  |
|  | Polycystic ovary syndrome | 6 (33.3%) |  |
|  | Endometriosis | 3 (16.7 %) |  |
|  | Oligomenorrhea | 2 (11.1%) |  |
| n. oocytes - pick-up |  | 127 |  |
| Oocytes metaphase I | 10 (7.9%) |
| Oocytes metaphase II | 117 (92.1%) |
| Fertilized oocytes | 117 (92.1%) |
| Blastocyst | 107 (91.5%) |
| Transfers performed | 17 (94.4%) |
| Beta HCG > 400 UI/mL | 8 (47.1%) |
| Miscarriages | 4 (23.5%) |
| Live birth | 4 (23.5%) |

IUI: Intra-uterine insemination; FIVET: In Vitro Fertilisation and Embryo-Transfer; ICSI: IntraCytoplasmic Sperm Injection.

### 2.2 Collection of biological samples

For follicular fluids sampling, BD glass tubes treated with reinforced USP Type III non-siliconized, 5 ml with red cap without additives were used and always stored horizontally. For the pick-up, Wallace 17G needles were used for all patients, in place of Falcon conical tubes in PPE, preheated BD glass tubes were used (Becton, Dickinson and Company, 2100 Derry Road West Mississauga, Ontario, Canada), follicular fluid was observed in preheated glass Petri dishes and then placed in BD glass tubes. All follicular fluids were processed to assess the possible presence of blood traces with the Albumin test, only suitable ones were stored at −20°C and analyzed for MPs.

### 2.3 Extraction and dosage of MPs

The samples of follicular fluid were prepared for extracting MPs according to a new patented method nationally and internationally protected (PCT/IB2019/051838 of March 7, 2019 coupled with the accepted Italian patent number 102018000003337 of March 07, 2018) validated with a recovery >81% (calculated using fortified samples with microparticles based on 3 μm red PS particles purchased from Merck – Sigma Aldrich, Germany, with a recovery ranging of 83–103%) and a LOD of 0.1 µm (Zuccarello et al., 2019a, 2019b). This methodological approach doesn’t use the filtration step to avoid the irremediable loss of NPs or MPs with size lowest to the pore filter diameter.

We identified and measured extracted MPs (as the sum of all types of plastics) focusing our results on MPs <10 µm (for better biological plausibility of their occurrence in this biological fluid against MPs with the highest sizes). Qualitative and quantitative determinations were performed using a Zeiss Scanning Electron Microscopy (SEM Zeiss LEO-1430) coupled with an EDX (X Energy Dispersion Detector) detector using the AZTEC Version 5.1 software. The EDX use permitted us to discriminate between plastic and not-plastic particles. The calculation was applied to an overall reading area within 1 mm^2^ of stub (Aluminium-Copper stub, coated with pure gold), examining 228 fields at 1500 X magnification. Results were expressed as the number of particles per ml (p/ml) of follicular fluid sample.

To prevent potential cross-samples or environmental sample contamination from personal synthetic clothing (nylon, polyester, etc…) and plastics use during the process of sample preparation, authors adopted the following preventive measures: glassware and metal equipment were used whenever possible thoroughly rinsed with filtered (1 µm - Whatman glass filter) Ultra-Performance Liquid Chromatography-Mass Spectrometry (UPLC-MS) Grade water and acetone; dust-free nitrile gloves were worn, use only of clean 100% cotton-based laboratory coats; all samples were extracted in a clean room under a horizontal laminar flow cabinet with controlled access; sample containers were preserved with glass box and protected by aluminium foil when not handled. Three reagent blanks were run with each batch of samples, and any measurable MPs weren’t detected in these blanks, showing that any potential contamination occurring from the sample treatment was null.

### 2.4 Dosage of biochemical markers

To determine the concentrations of hFSH, hLH, and Estradiol, an immunoassay in chemiluminescence with paramagnetic particles was used, using the Access 2 Immunoassay System by Beckman Coulter according to the manufacturer’s instructions. The values are expressed as follows: hLH and hFSH in mIU/mL; Estradiol in pg/mL. For the quantitative measurement of circulating anti-Müllerian Hormone the ELFA (Enzyme Linked Fluorescent Assay) technique was used, with an automatic VIDAS analyzer from BioMerieux-France, according to the manufacturer’s instructions. Values are expressed in ng/mL.

### 2.5 Statistical analysis

A statistical descriptive analysis was carried out using the Excel 2021 (Microsoft, Excel 2021). A preliminary explorative analysis of data suggested to authors the application of Pearson’s correlation test to evaluate the positive, negative, or null linear correlation between the pair of variables. For each pair of variables, Pearson’s product moment correlation coefficient “*r”* was calculated as null, weak, moderate and straight force of linear correlation approaching the Interpretation data suggested by (Chen and Anderson, 2023). The interpretation of the *r* value (https://www.ncl.ac.uk/webtemplate/ask-assets/external/maths-resources/statistics/regression-and-correlation/strength-of-correlation.html) is reported in the supporting material.

The significance level was set at *p-value* = 0.05. A *p-value* > 0.05 means that deviation from the null hypothesis is not statistically significant and the null hypothesis is not rejected (Andrade, 2019).

## 3. Results and discussion

In the present study, microplastics (dimensions <10 m) were detected in human follicular fluid in 14 out of 18 participants, with an average of 2191 p/ml (0 - 7181p/ml) and with a mean diameter of MPs of 4.48 (3.18-5.54 m) (see Tab. 1). Results for all samples showing the details are reported in Table 2 and Figure 1. These latter shows that the total concentration of MPs in sample N. 18 is the highest, followed by samples No. 4 and No. 7. No correlation was found between microplastic concentration, fertilization, miscarriages, and live birth. Instead, a moderate correlation was found between MPs <10 µm and FSH (*r* = 0.52) with a *p*-value <0.05. In addition, weak correlations between MPs <10µm with BMI (*r* = 0.31), age (*r* = 0.24) and E2 (*r* = 0.22) but all with *p* >0.05, were found (See Supporting material). Probably, the weak correlations could be justified by the limited number of analyzed samples.

**Figure 1.**
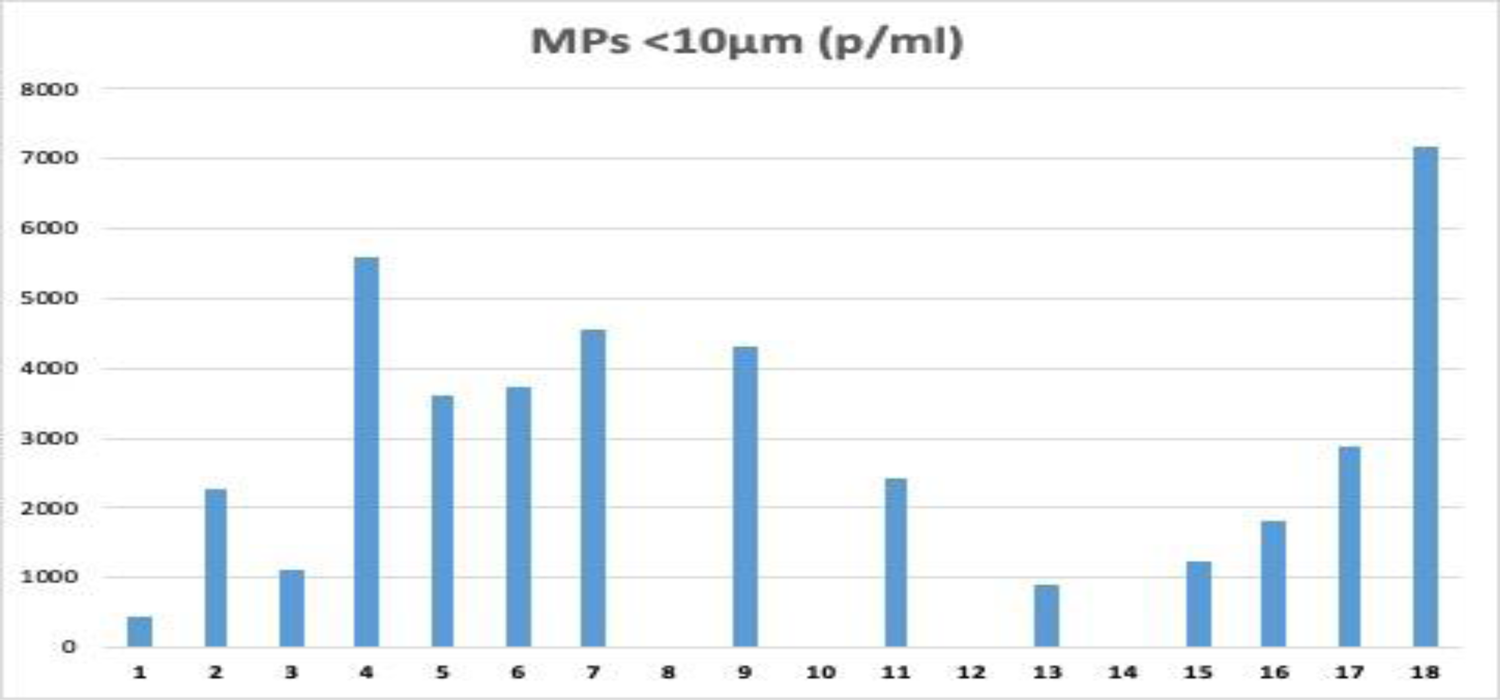
Concentration of MPs in follicular fluid samples.

**Table 2.** Results of MPs in the follicular fluid samples.

| ID samples | MPs <10µm (p/ml) | Average diameter µm (particles <10 µm) |
| --- | --- | --- |
| 1 | 442 | 4.12 |
| 2 | 2276 | 4.78 |
| 3 | 1101 | 4.58 |
| 4 | 5600 | 4.94 |
| 5 | 3601 | 3.9 |
| 6 | 3736 | 5.5 |
| 7 | 4566 | 3.66 |
| 8 | 0 |  |
| 9 | 4326 | 4.76 |
| 10 | 0 |  |
| 11 | 2431 | 5.44 |
| 12 | 0 |  |
| 13 | 897 | 5.44 |
| 14 | 0 |  |
| 15 | 1242 | 5.28 |
| 15 | 1816 | 5.22 |
| 17 | 2875 | 5.32 |
| 18 | 7181 | 4.5 |
| min | 0 | 3.66 |
| max | 7181 | 5.54 |
| average | 2196 | 4.46 |

Scanning Electron Microscope (SEM) is a useful tool in providing high-resolution particle surface structure characteristics of the material. In this study, a field emission scanning electron microscope was used to visualize microplastics and evaluate the size and shape of the particles in human follicular fluid samples. Figure 2 shows the selected SEM images of typical microplastics isolated from human ovarian follicular fluid.

**Figure 2.**
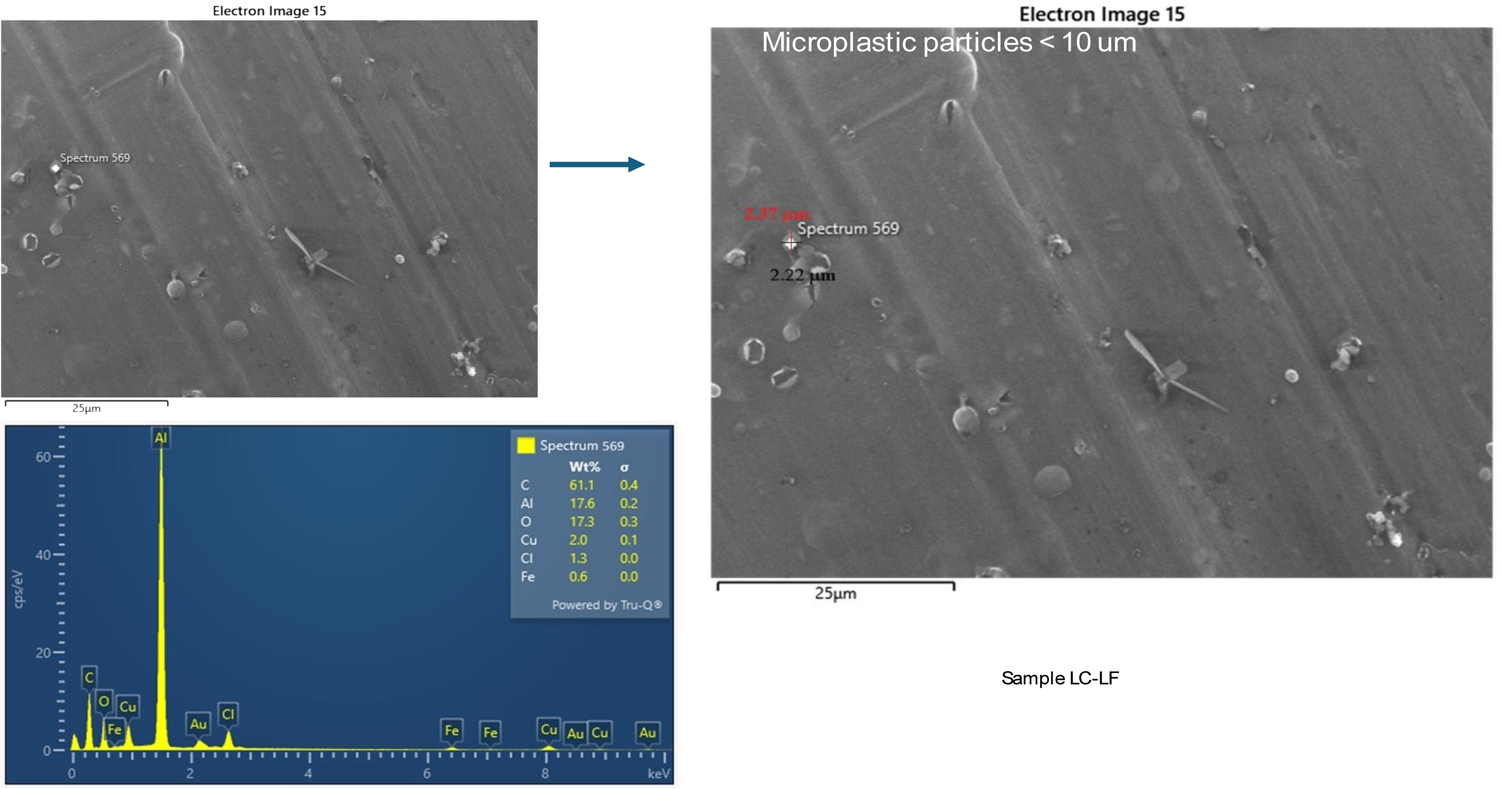
Selected SEM-EDX image microplastics in follicular fluid.

## 4. Discussion

The usage behaviors of plastic products of the participants are very similar since all of them declared to use plastic-made items in their life, therefore trying to understand and differentiate a possible exposure route is hardly truthful. We can only speculate that micro and nano plastics enter the human body by ingestion and/or inhalation and direct skin contact as now globally recognized (Pironti et al., 2021).

Thousands of micro and nano plastic particles can be accumulated by adults during their lifetime (Lim, 2021), as they have been found in various human tissues, including kidneys, liver, hair, lungs, and spleen (Kutralam-Muniasamy et al., 2023) but also in meconium, breast milk, placenta, blood (Leslie et al., 2022), urine (Pironti et al., 2023) and sperm (Montano et al., 2023). In literature is reported that MNP exposure was associated with microstructural changes in the reproductive system of female animals, such as oviduct dilatation, an increase in the number of ovarian cysts and corpus luteum, a thinner granular layer of secondary follicles, and a decrease in the number of developing follicles (An et al., 2021; Hou et al., 2021; Park et al., 2020). A potential mechanism that may explain the presence of MPs in the human ovarian follicular fluid could be related to the possibility that MPs pass from the bloodstream to the follicle and thus cross the blood-follicle barrier, which is a more dynamic and less stable barrier than the blood-testicular barrier, perhaps closer to the placental barrier. Therefore, microplastics could enter the ovaries through the circulatory system reaching the granulosa cells (Figure 3). The biggest cells in the follicle, ovarian granulosa cells (GCs), are responsible for secreting estrogen and progesterone. GC proliferation and differentiation impact all major ovarian functional processes, including follicular growth and development, ovulation, luteal formation, and steroid hormone production. MPs have been demonstrated to enter rat ovarian GCs, resulting in a substantial decrease in blood levels of anti-Mullerian hormone (AMH) and E2 expression, as well as enhanced expression of fibrosis markers (An et al., 2021). This ultimately led to sluggish follicular development and an irregular estrous cycle. In addition, it has been reported that PS-MPs lowered cytoskeletal protein levels in rat ovaries, including α-microtubulin, breaking physical-functional linkages between the ovarian stroma and parenchyma and compromising oocyte-cell synchronization (Haddadi et al., 2022). This work also reported on the location and accumulation of PS-MPs in the rat ovary (Haddadi et al., 2022).

**Figure 3.**
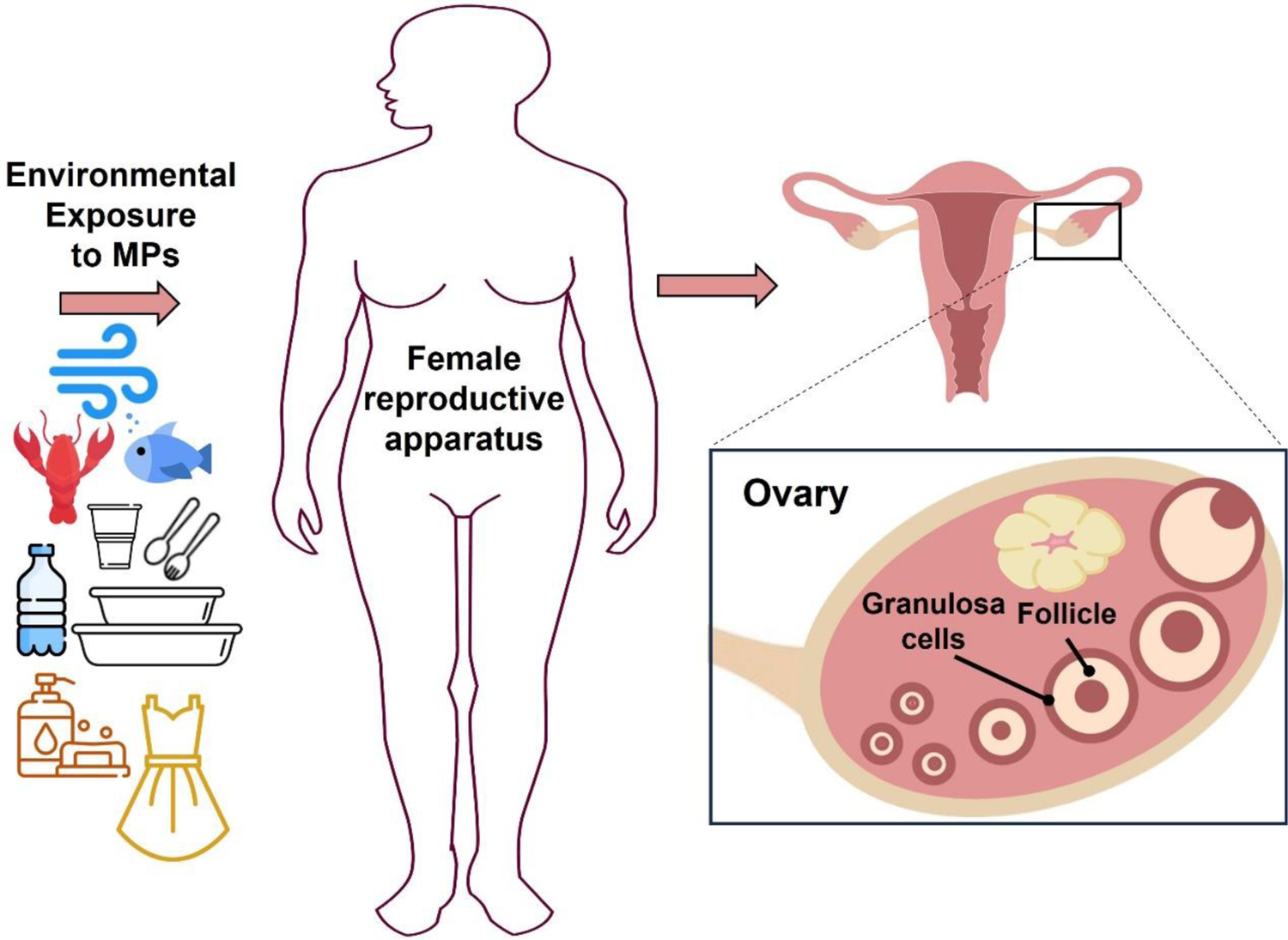
Schematization of the mechanism by which MPs pass into the ovarian follicular fluid: through environmental exposure (inhalation, ingestion and dermal contact) they enter the human body, reaching the female reproductive apparatus, particularly crossing the blood-follicle barrier.

Tests done in rodents reveal that MPs reach the vaginal canal and appear to induce a reduction in follicle number while boosting ovarian oxidative stress and apoptosis in granulosa cells. In addition, maternal exposure to MPs enhanced oxidative stress in oocytes while decreasing polar body extrusion. Oral administration of MPs in gravid mice resulted in a faster rate of embryonic resorption and placental and fetal development (Yang, 2023). Moreover, MPs may alter oocyte maturation and function, resulting in decreased fertility (Zhang et al., 2023). In this work, the authors demonstrated that the administration of PS-MPs impaired oocyte maturation and decreased the quality of oocytes by promoting apoptosis via increased ROS levels. Furthermore, in combination with DEHP, PS-MPs synergistically activated the CNR1/CRBN/YY1/CYP2E1 and Hippo pathways and induced ovarian granulosa cell cycle arrest and necroptosis via the generation of OS and DNA oxidative damage (Wu et al., 2023). In summary, exposure to MPs may exacerbate several reproductive toxicities in female mammals, mainly through OS, apoptosis and fibrosis. These mechanisms may ultimately lead to a range of structural and functional reproductive alterations, including reduced oocyte numbers, impaired follicular growth, granulosa cell apoptosis, reduced ovarian reserve function, and uterine and ovarian fibrosis. After all, studies have shown that the accumulation of ROS can lead to apoptosis of GCs and cause follicular atresia in mice (Shen et al., 2012), which may be the causal factor resulting in anovulatory infertility (Liu et al., 2019; Wang et al., 2018). Wei et al. (Wei et al., 2022) exposed male and female mice to fluorescent PS-MPs of 5.0–5.9 μm diameter, for two days and found that fluorescent PS-MPs had entered the testes and ovaries of the mice being more fluorescent PS-MPs in the ovaries than in the testes. Moreover, they found that the ovary size in the PS-MPs treatment group was smaller and the number of follicles in the PS-MPs treatment groups at each stage was lower than that of the control group.

The ovary is an important reproductive and endocrine organ that produces oocytes and secretes steroid hormones. It is particularly prone to be affected by many environmental substances. A recent study found that MPs accumulated in rats’ ovaries, inducing granulosa cell apoptosis through oxidative stress and fibrosis of the ovary via Wnt/β-Catenin signaling pathway activation (An et al., 2021) and causing abnormal folliculogenesis (Haddadi et al., 2022). The follicle is the basic structural unit of the ovary, which can be divided into preantral follicles and antral follicles. During the growth of the oocyte, the follicle is filled with follicular fluid, which plays a crucial role in the maturation and development of the oocyte. Haddadi et al. (Haddadi et al., 2022) assessed the impact of oral exposure, during four estrous cycles, of 5 μm PS-MPs on ovarian function in rats. They found particles of PS-MPs in the duodenum and the different compartments of the ovarian tissue. A reduced relative ovarian weights and a reduced serum concentration of estradiol were associated with the toxicity of PS-MPs, causing also an alteration in the folliculogenesis and the estrous cycle duration. These defective ovarian functions are most probably caused by the induction of oxidative stress, evidenced by increased superoxide dismutase (SOD) and catalase (CAT) activities and an increased malondialdehyde (MDA) concentration as well as a decreased protein sulfhydryl (PSH) level in the rat ovary. Moreover, they demonstrated a significant decrease in the expression of cytoskeletal proteins by immunofluorescence and RT-PCR: α-tubulin and disheveled-associated activator of morphogenesis (DAAM-1) in the ovary of rats exposed to PS-MPs at proteomic and transcriptomic levels.

Regarding the correlation between MPs concentration and FSH, an interesting study on mice was focused on examining the various impacts of PS-MPs on the reproductive systems of both female and male mice, as well as assessing the impact of PS-MPs exposure on their fertility. Results indicated that PS-MPs led to greater accumulation and oxidative stress in the ovary compared to the testis, reduced ovary size and follicle number, increased FSH levels (Wei et al., 2022).

As for the weak correlation found between MPs concentration and BMI, in current literature the “obesogen” effects of these particles on cells are well described, in fact, Kannan & Vimalkumar (Kannan and Vimalkumar, 2021) reported this phenomenon in kidney and liver cells for MPs <20 µm, in which MPs altered energy and fatty acid metabolism that can affect, in the end, body weight. In mice the obesogenic role of MPs was explained in perturbing the gut-liver-adipose axis and altering nuclear receptor signaling and intermediary metabolism by Zhao et al., (Zhao et al., 2024). Jeong et al. demonstrated, through omic analyses, that environmental NPs can act as obesogens in childhood (Jeong et al., 2024). In the Najahi et al. study (Najahi et al., 2022), authors demonstrated that adipose mesenchymal stromal cells, when exposed to MPs <1 um, these do not differentiate in the normal programmed tissue. Additionally, there may also be some effect on E2, considering that microplastics have been shown to have a negative effect on granulosa cells, which produce this estrogen (An et al., 2021). Of course, these evaluations are based on still limited numbers, so more extensive recruitment is necessary to have more reliable elaborations on the effects of these emerging contaminants on the reproductive function of human females (Geng et al., 2023; Zurub et al., 2024).

## 5. Conclusion

Once again plastic molecules were found to invade our bodies at deeper levels. As far as we know, this is the first study to provide evidence for microplastics’ presence in ovarian follicular fluid in women undergoing assisted reproductive treatment. Although a certain relationship between quantities of MPs and some important parameters for the regulation of ovarian function has been observed in this study, albeit needing to be evaluated with more consistent numbers, it indicates the need to continue in this direction to better understand the effects of these emerging contaminants on female reproductive health. Therefore, this study should be considered extremely relevant for the scientific community.

## CRediT authorship contribution statement

Luigi Montano: Conceptualization; project administration; writing of the original draft; Writing - review & editing; Supervision. Salvatore Raimondo: investigation, data curation, resources, Writing - review & editing. Marina Piscopo: writing of the original draft, Writing - review & editing. Maria Ricciardi: writing of the original draft; Writing - review & editing. Antonino Guglielmino: review & editing, visualization. Sandrine Chamayou: review & editing; visualization. Raffaella Gentile; visualization. Mariacira Gentile: visualization. Paola Rapisarda: investigations; writing of the original draft; Writing-review & editing. Gea Oliveri Conti: investigations; writing of the original draft; Writing-review & editing. Margherita Ferrante: conceptualization; resources, data curation, software, validation, formal analysis; writing of the original draft; Writing-review & editing. Oriana Motta: Conceptualization; resources, writing of the original draft; Writing-review & editing; Supervision.

## Institutional Review Board Statement

Informed consent was obtained from all subjects involved in the study. The study was conducted under the Declaration of Helsinki and the Italian Code regarding the protection of personal data (Legislative Decree 196/2003) and falls within the scope of the EcoFoodFertility project (https://www.ecofoodfertility.it, accessed on 08 Febr 2024), approved by the Ethical Committee of the Local Health Authority Campania Sud-Salerno (Committee code n. 43 of 30 June 2015) Italy; the participants were informed about the general purpose of the research, the anonymity of the answers, and the voluntary nature of participation, and they signed informed consent. There were no incentives given.

## Informed Consent Statement

Informed consent was obtained from all subjects involved in the study.

## Data Availability

All data produced in the present work are contained in the manuscript

## Acknowledgment

This study forms part of the EcoFoodFertility project, which is a multicenter biomonitoring study to develop a better understanding of the environmental impact of toxicants on human health, especially reproductive health.

## Supporting materials

### Additional information on study participants

**Table S1.** Detailed information on all participant parameters.

| ID<br>sam<br>ples | MPs<br><10µm<br>(p/ml) | average<br>diameter<br>µm<br>(particles<br><10 µm) | age | BMI | waist | waist-to-hip<br>ratio | nullipar<br>ous | multiparo<br>us | contrace<br>ptives | previous assisted<br>reproduction procedures |  |  |
| --- | --- | --- | --- | --- | --- | --- | --- | --- | --- | --- | --- | --- |
|  |  |  |  |  |  |  |  |  |  | IUI | FIVET | ICSI |
| 1 | 442 | 4.12 | 31-35 | 23.4 | 85 | 0.73 | yes | no | yes | 1 | 0 | 2 |
| 2 | 2276 | 4.78 | 31-35 | 30.2 | 88 | 0.92 | no | 1 | yes | 0 | 0 | 1 |
| 3 | 1101 | 4.58 | 25-30 | 25.0 | 70 | 0.55 | yes | no | yes | 0 | 0 | 0 |
| 4 | 5600 | 4.94 | 35-40 | 33 | 80 | 0.9 | yes | no | yes | 0 | 0 | 0 |
| 5 | 3601 | 3.9 | 35-40 | 38 | 110 | 0.95 | yes | no | yes | 2 | 0 | 0 |
| 6 | 3736 | 5.5 | 40-45 | 27 | 105 | 0.5 | yes | no | yes | 3 | 1 | 0 |
| 7 | 4566 | 3.66 | 31-35 | 23 | 71 | 0.78 | yes | no | no | 0 | 0 | 0 |
| 8 | 0 |  | 31-35 | 25 | 96 | 0.8 | yes | no | no | 3 | 0 | 1 |
| 9 | 4326 | 4.76 | 40-45 | 24 | 74 | 0.68 | no | 1 | no | 0 | 0 | 0 |
| 10 | 0 |  | 35-40 | 27 | 70 | 0.68 | yes | no | no | 3 | 0 | 0 |
| 11 | 2431 | 5.44 | 35-40 | 23 | 69 | 0.7 | yes | no | no | 0 | 0 | 1 |
| 12 | 0 |  | 35-40 | 29 | 81 | 0.69 | yes | no | yes | 0 | 0 | 1 |
| 13 | 897 | 5.44 | 25-30 | 28.2 | 76 | 0.71 | no | 1 | yes | 0 | 0 | 0 |
| 14 | 0 |  | 25-30 | 25.6 | 77 | 0.7 | yes | no | yes | 0 | 0 | 0 |
| 15 | 1242 | 5.28 | 40-45 | 26.2 | 87 | 0.71 | no | 2 | yes | 0 | 0 | 0 |
| 15 | 1816 | 5.22 | 40-45 | 20.7 | 80 | 0.7 | yes | no | no | 0 | 0 | 0 |
| 17 | 2875 | 5.32 | 25-30 | 27.1 | 80 | 0.81 | yes | no | no | 0 | 0 | 0 |
| 18 | 7181 | 4.5 | 35-40 | 29.6 | 87.2 | 0.86 | yes | no | yes | 1 | 0 | 1 |

**Table S2.**
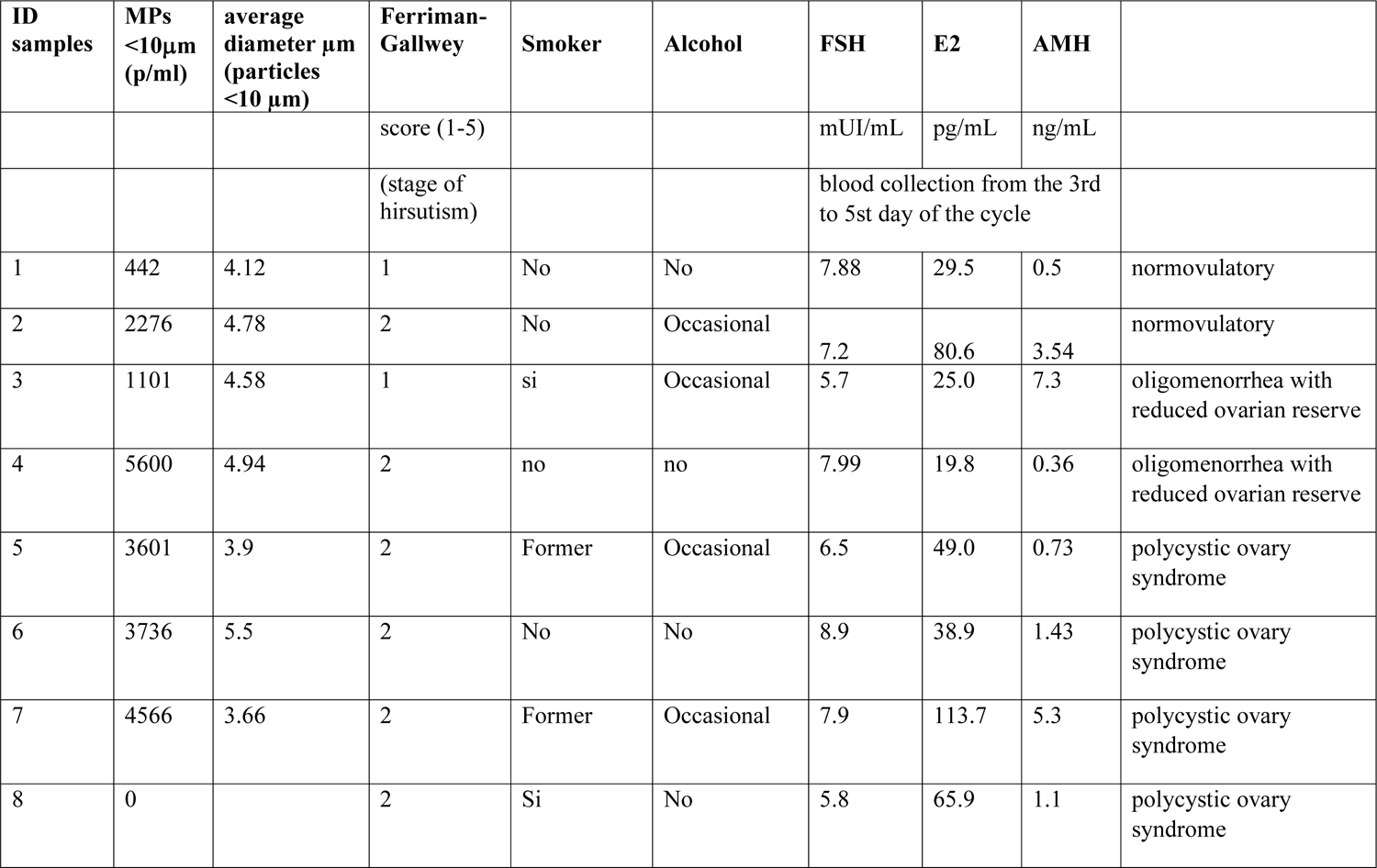
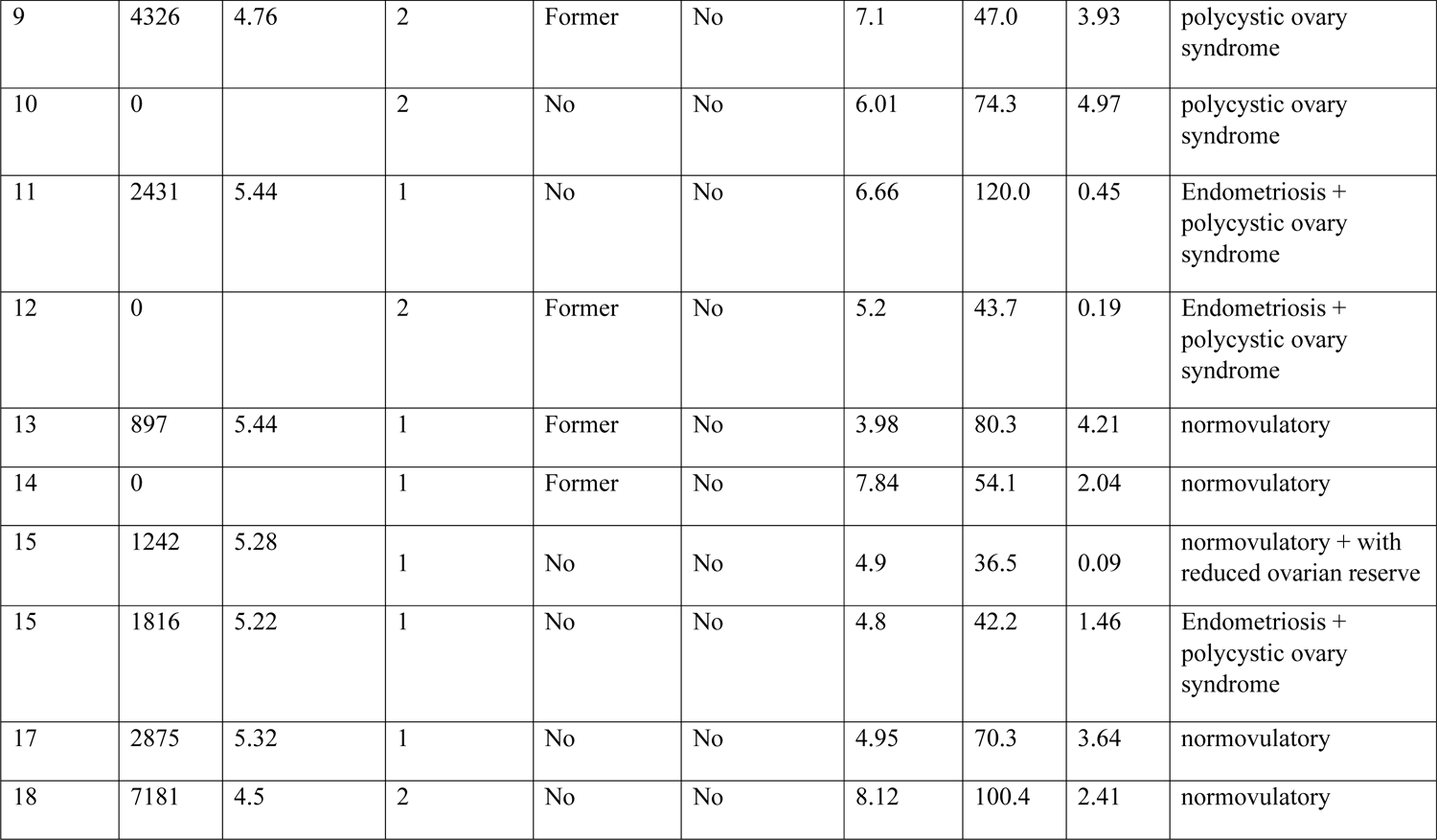
Detailed information on all participant parameters (continued).

| ID samples | MPs <10µm (p/ml) | average diameter µm (particles <10 µm) | Ferriman-Gallwey | Smoker | Alcohol | FSH | E2 | AMH |  |
| --- | --- | --- | --- | --- | --- | --- | --- | --- | --- |
|  |  |  | score (1-5) |  |  | mUI/mL | pg/mL | ng/mL |  |
|  |  |  | (stage of hirsutism) |  |  | blood collection from the 3rd to 5st day of the cycle |  |  |  |
| 1 | 442 | 4.12 | 1 | No | No | 7.88 | 29.5 | 0.5 | normovulatory |
| 2 | 2276 | 4.78 | 2 | No | Occasional | 7.2 | 80.6 | 3.54 | normovulatory |
| 3 | 1101 | 4.58 | 1 | si | Occasional | 5.7 | 25.0 | 7.3 | oligomenorrhea with reduced ovarian reserve |
| 4 | 5600 | 4.94 | 2 | no | no | 7.99 | 19.8 | 0.36 | oligomenorrhea with reduced ovarian reserve |
| 5 | 3601 | 3.9 | 2 | Former | Occasional | 6.5 | 49.0 | 0.73 | polycystic ovary syndrome |
| 6 | 3736 | 5.5 | 2 | No | No | 8.9 | 38.9 | 1.43 | polycystic ovary syndrome |
| 7 | 4566 | 3.66 | 2 | Former | Occasional | 7.9 | 113.7 | 5.3 | polycystic ovary syndrome |
| 8 | 0 |  | 2 | Si | No | 5.8 | 65.9 | 1.1 | polycystic ovary syndrome |
| 9 | 4326 | 4.76 | 2 | Former | No | 7.1 | 47.0 | 3.93 | polycystic ovary syndrome |
| 10 | 0 |  | 2 | No | No | 6.01 | 74.3 | 4.97 | polycystic ovary syndrome |
| 11 | 2431 | 5.44 | 1 | No | No | 6.66 | 120.0 | 0.45 | Endometriosis + polycystic ovary syndrome |
| 12 | 0 |  | 2 | Former | No | 5.2 | 43.7 | 0.19 | Endometriosis + polycystic ovary syndrome |
| 13 | 897 | 5.44 | 1 | Former | No | 3.98 | 80.3 | 4.21 | normovulatory |
| 14 | 0 |  | 1 | Former | No | 7.84 | 54.1 | 2.04 | normovulatory |
| 15 | 1242 | 5.28 | 1 | No | No | 4.9 | 36.5 | 0.09 | normovulatory + with reduced ovarian reserve |
| 15 | 1816 | 5.22 | 1 | No | No | 4.8 | 42.2 | 1.46 | Endometriosis + polycystic ovary syndrome |
| 17 | 2875 | 5.32 | 1 | No | No | 4.95 | 70.3 | 3.64 | normovulatory |
| 18 | 7181 | 4.5 | 2 | No | No | 8.12 | 100.4 | 2.41 | normovulatory |

**Table S3.** Detailed information on all participant parameters (continued).

| ID samples | MPs <10µm (p/ml) | average diameter µm (particles <10 µm) | drug therapy | retrived ovocytes | Metaphase | Metaphase | ferti lized oocy tes | eight-cell embryos | transfer perform ed | beta HCG > 400 UI/ml | aborti ons | live birth |
| --- | --- | --- | --- | --- | --- | --- | --- | --- | --- | --- | --- | --- |
|  |  |  |  | Pick-up | oocytes I | oocytes II |  |  |  |  |  |  |
| 1 | 442 | 4.12 |  | 6 | 0 | 6 | 6 | 6 | yes | negative | / | / |
| 2 | 2276 | 4.78 |  | 7 | 0 | 7 | 7 | 7 | yes | positive | / | yes |
| 3 | 1101 | 4.58 |  | 7 | 0 | 7 | 7 | 5 | yes | negative | / | / |
| 4 | 5600 | 4.94 |  | 7 | 0 | 7 | 7 | 6 | yes | positive | / | yes |
| 5 | 3601 | 3.9 |  | 7 | 1 | 6 | 7 | 6 | yes | positive | yes | / |
| 6 | 3736 | 5.5 |  | 8 | 2 | 6 | 7 | 7 | yes | positive | yes | / |
| 7 | 4566 | 3.66 |  | 5 | 1 | 4 | 4 | 1 | no | negative | / | / |
| 8 | 0 |  |  | 7 | 0 | 7 | 7 | 7 | yes | negative | / | / |
| 9 | 4326 | 4.76 |  | 7 | 0 | 7 | 7 | 7 | yes | negative | / | / |
| 10 | 0 |  |  | 8 | 0 | 8 | 8 | 8 | yes | negative | / | / |
| 11 | 2431 | 5.44 |  | 4 | 0 | 4 | 4 | 4 | yes | negative | / | / |
| 12 | 0 |  |  | 9 | 3 | 6 | 6 | 6 | yes | positive | / | yes |
| 13 | 897 | 5.44 |  | 11 | 2 | 9 | 9 | 9 | yes | negative | / | / |
| 14 | 0 |  | betametha sone | 5 | 0 | 5 | 5 | 4 | yes | negative | / | / |
| 15 | 1242 | 5.28 | betametha<br>sone | 7 | 0 | 7 | 6 | 5 | yes | positive | yes | / |
| 15 | 1816 | 5.22 |  | 7 | 1 | 6 | 6 | 6 | yes | positive | / | yes |
| 17 | 2875 | 5.32 |  | 9 | 0 | 9 | 8 | 7 | yes | negative | / | / |
| 18 | 7181 | 4.5 |  | 6 | 0 | 6 | 6 | 6 | yes | positive | yes | / |

### Statistical analyses

**Table S4.**
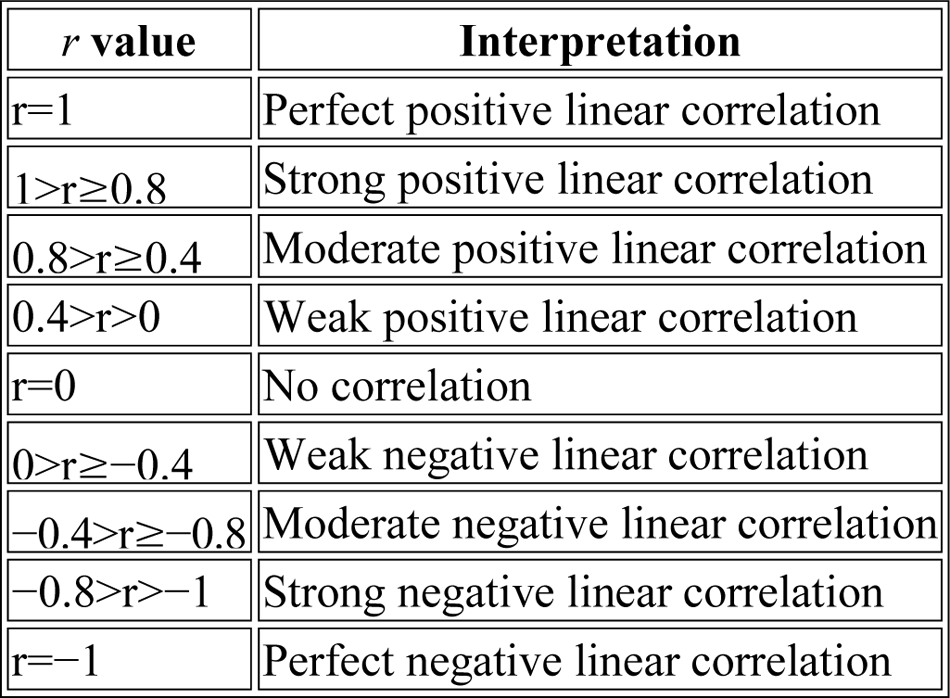
Interpretation of the *r* value.

| *r* value | Interpretation |
| --- | --- |
| $r=1$ | Perfect positive linear correlation |
| $1>r\geq 0.8$ | Strong positive linear correlation |
| $0.8>r\geq 0.4$ | Moderate positive linear correlation |
| $0.4>r>0$ | Weak positive linear correlation |
| $r=0$ | No correlation |
| $0>r\geq -0.4$ | Weak negative linear correlation |
| $-0.4>r\geq -0.8$ | Moderate negative linear correlation |
| $-0.8>r>-1$ | Strong negative linear correlation |
| $r=-1$ | Perfect negative linear correlation |

**Figure S1.**
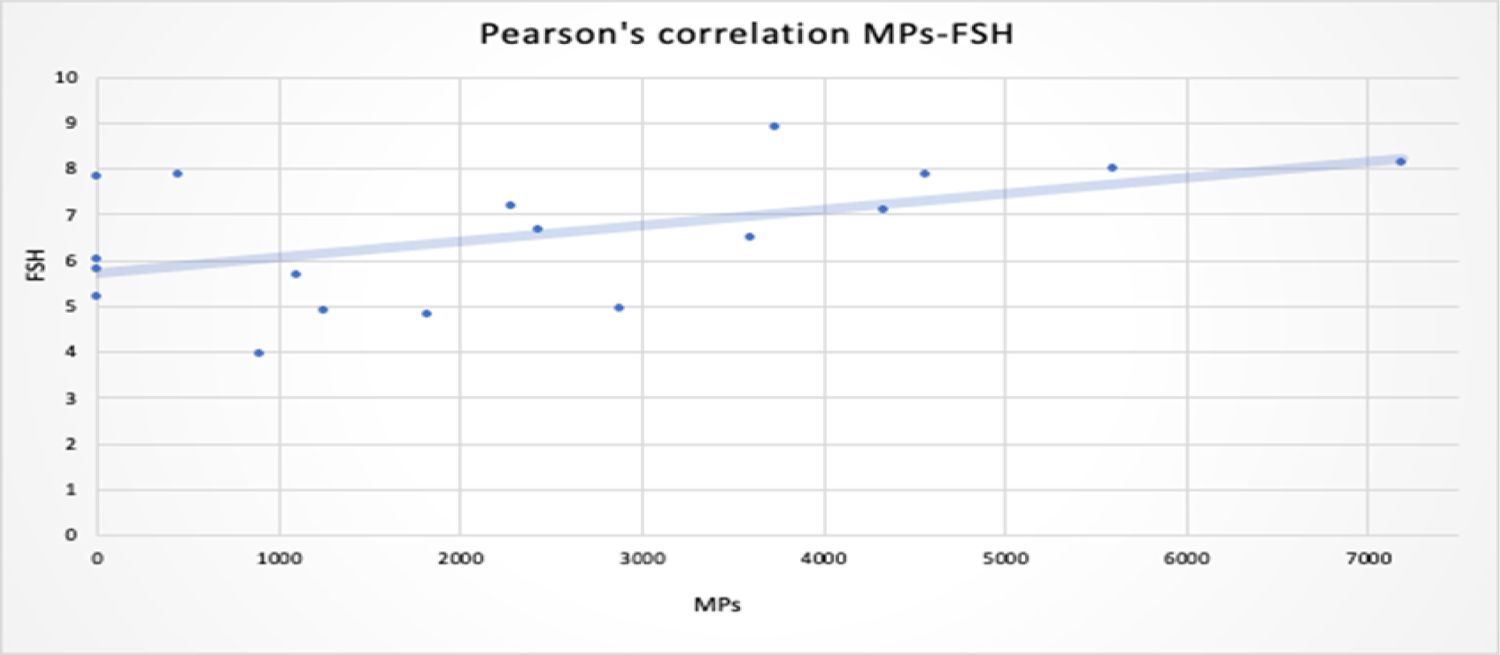
Correlation graph between number of MPs and FSH.

**Figure S2.**
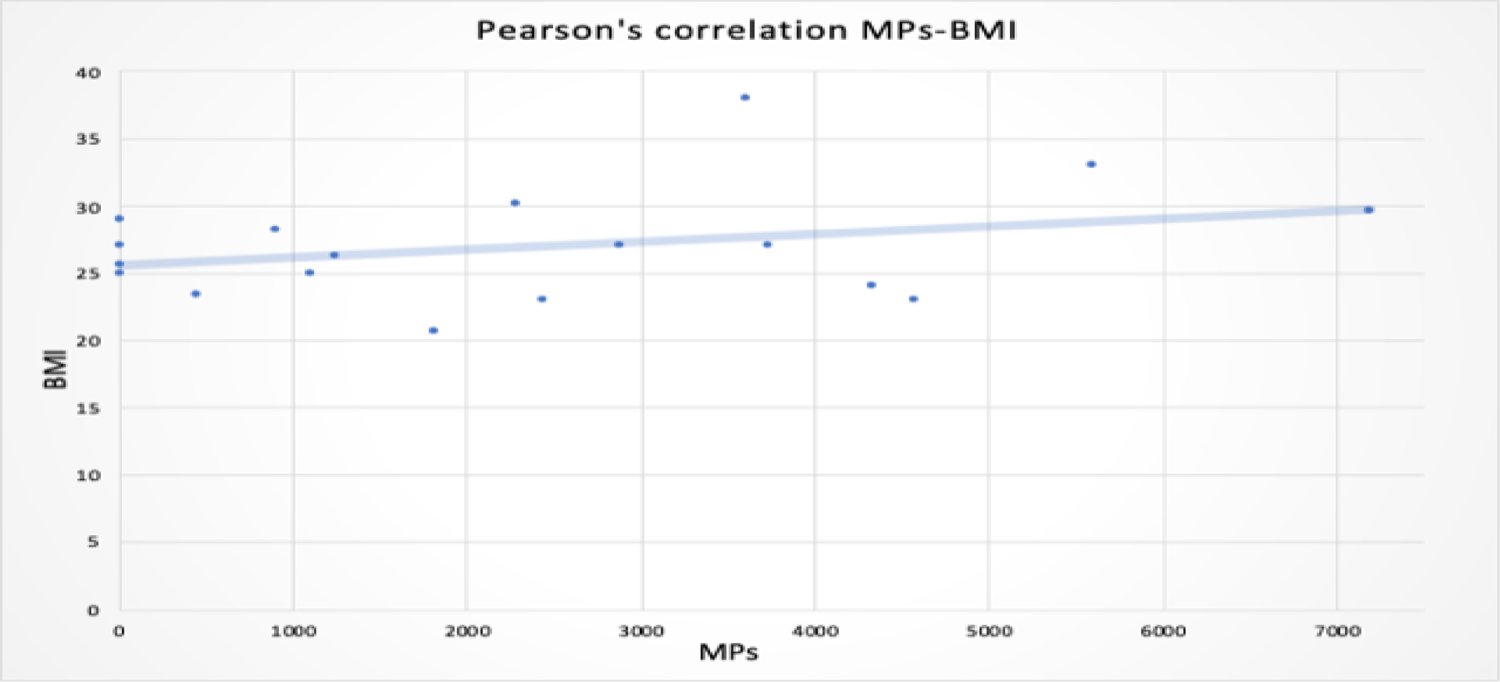
Correlation graph between number of MPs and BMI.

**Figure S3.**
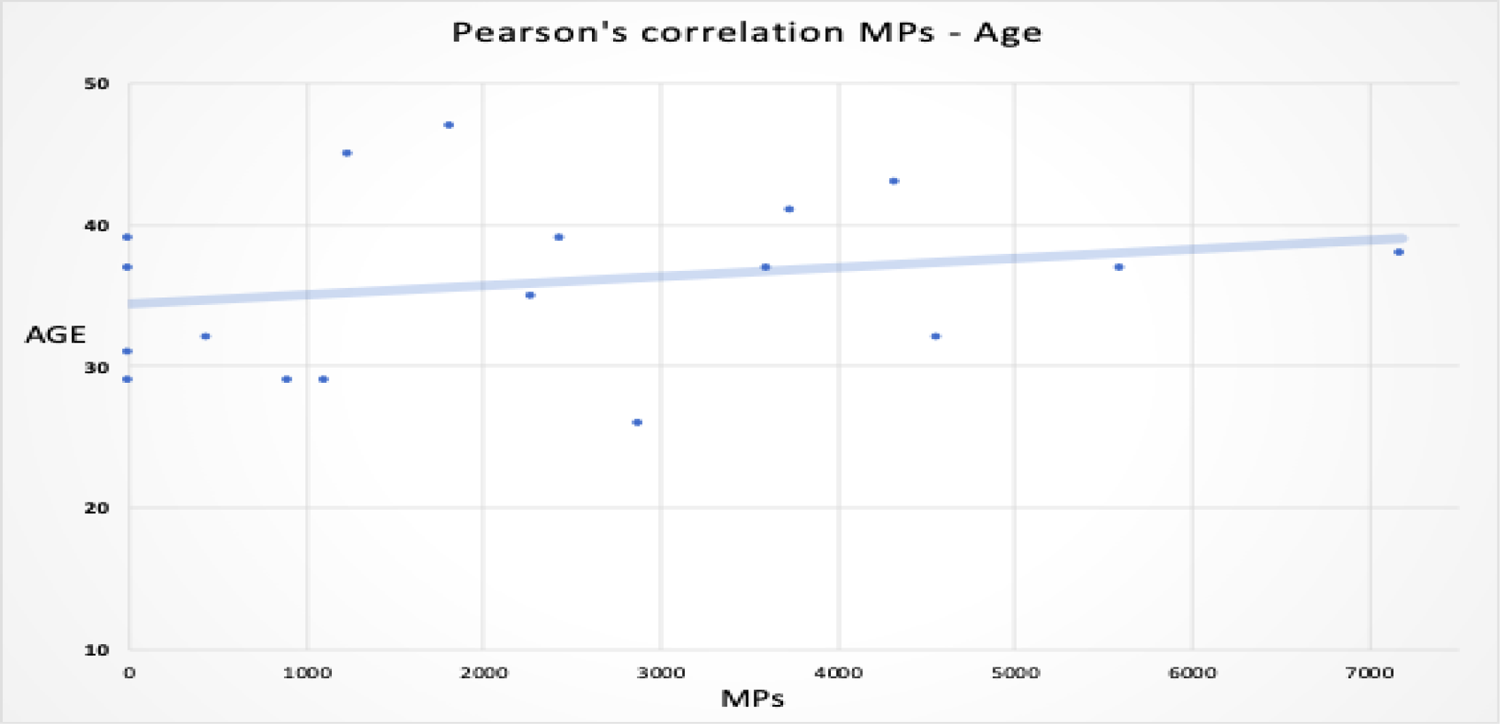
Correlation graph between number of MPs and Age.

**Figure S4.**
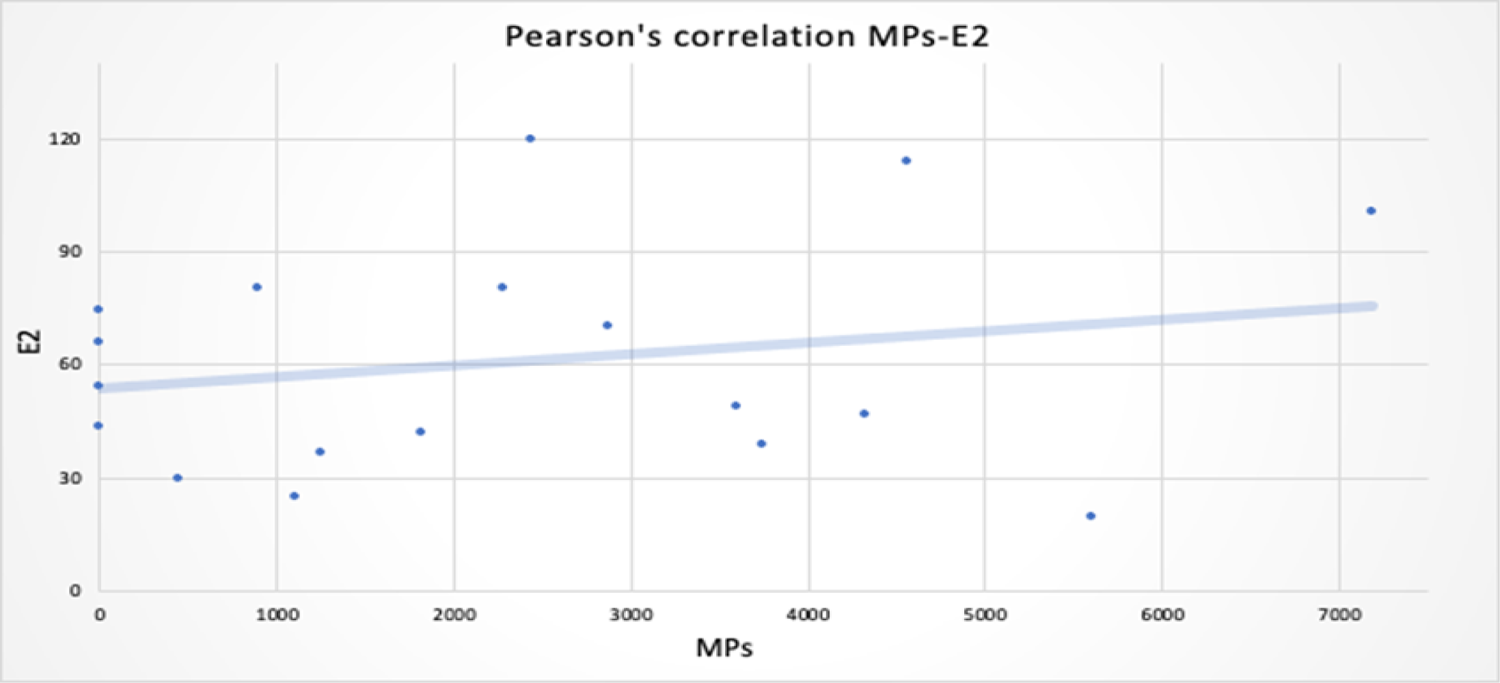
Correlation graph between number of MPs and E2.

## References

Aitken, R.J., 2022. The changing tide of human fertility. Human Reproduction 37, 629–638. 10.1093/humrep/deac011

An, R., Wang, X., Yang, L., Zhang, J., Wang, N., Xu, F., Hou, Y., Zhang, H., Zhang, L., 2021. Polystyrene microplastics cause granulosa cells apoptosis and fibrosis in ovary through oxidative stress in rats. Toxicology 449, 152665. 10.1016/j.tox.2020.152665

Andrade, C., 2019. The P Value and Statistical Significance: Misunderstandings, Explanations, Challenges, and Alternatives. Indian Journal of Psychological Medicine 41, 210–215. 10.4103/IJPSYM.IJPSYM_193_19

Bergamo, P., Volpe, M.G., Lorenzetti, S., Mantovani, A., Notari, T., Cocca, E., Cerullo, S., Di Stasio, M., Cerino, P., Montano, L., 2016. Human semen as an early, sensitive biomarker of highly polluted living environment in healthy men: A pilot biomonitoring study on trace elements in blood and semen and their relationship with sperm quality and RedOx status. Reproductive Toxicology 66, 1–9. 10.1016/j.reprotox.2016.07.018

Chen, D., Anderson, C.J., 2023. Categorical data analysis, in: Tierney, R.J., Rizvi, F., Ercikan, K. (Eds.), International Encyclopedia of Education (Fourth Edition). Elsevier, Oxford, pp. 575–582. 10.1016/B978-0-12-818630-5.10070-3

Coffin, S., Bouwmeester, H., Brander, S., Damdimopoulou, P., Gouin, T., Hermabessiere, L., Khan, E., Koelmans, A.A., Lemieux, C.L., Teerds, K., Wagner, M., Weisberg, S.B., Wright, S., 2022. Development and application of a health-based framework for informing regulatory action in relation to exposure of microplastic particles in California drinking water. Microplastics and Nanoplastics 2, 12. 10.1186/s43591-022-00030-6

Ding, T., Yan, W., Zhou, T., Shen, W., Wang, T., Li, M., Zhou, S., Wu, M., Dai, J., Huang, K., Zhang, J., Chang, J., Wang, S., 2022. Endocrine disrupting chemicals impact on ovarian aging: Evidence from epidemiological and experimental evidence. Environmental Pollution 305, 119269. 10.1016/j.envpol.2022.119269

Ferrante, M., Pietro, Z., Allegui, C., Maria, F., Antonio, C., Pulvirenti, E., Favara, C., Chiara, C., Grasso, A., Omayma, M., Gea, O.C., Banni, M., 2022. Microplastics in fillets of Mediterranean seafood. A risk assessment study. Environmental Research 204, 112247. 10.1016/j.envres.2021.112247

Ferrero, G., Festa, R., Follia, L., Lettieri, G., Tarallo, S., Notari, T., Giarra, A., Marinaro, C., Pardini, B., Marano, A., Piaggeschi, G., Di Battista, C., Trifuoggi, M., Piscopo, M., Montano, L., Naccarati, A., 2024. Small noncoding RNAs and sperm nuclear basic proteins reflect the environmental impact on germ cells. Molecular Medicine 30, 12. 10.1186/s10020-023-00776-6

Gallo, A., Boni, R., Tosti, E., 2020. Gamete quality in a multistressor environment. Environment International 138, 105627. 10.1016/j.envint.2020.105627

Garrido Gamarro, E., 2022. Microplastics in food commodities: A food safety review on human exposure through dietary sources, Food Safety and Quality Series. FAO, Rome, Italy. 10.4060/cc2392en

Geng, Y., Liu, Z., Hu, R., Huang, Y., Li, F., Ma, W., Wu, X., Dong, H., Song, K., Xu, X., Zhang, Z., Song, Y., 2023. Toxicity of microplastics and nanoplastics: invisible killers of female fertility and offspring health. Front. Physiol. 14. 10.3389/fphys.2023.1254886

Gosden, R.G., Hunter, R.H., Telfer, E., Torrance, C., Brown, N., 1988. Physiological factors underlying the formation of ovarian follicular fluid. J Reprod Fertil 82, 813–825. 10.1530/jrf.0.0820813

Haddadi, A., Kessabi, K., Boughammoura, S., Rhouma, M.B., Mlouka, R., Banni, M., Messaoudi, I., 2022. Exposure to microplastics leads to a defective ovarian function and change in cytoskeleton protein expression in rat. Environ Sci Pollut Res 29, 34594–34606. 10.1007/s11356-021-18218-3

Hou, B., Wang, F., Liu, T., Wang, Z., 2021. Reproductive toxicity of polystyrene microplastics: In vivo experimental study on testicular toxicity in mice. Journal of Hazardous Materials 405, 124028. 10.1016/j.jhazmat.2020.124028

Huang, J., Zou, L., Bao, M., Feng, Q., Xia, W., Zhu, C., 2023. Toxicity of polystyrene nanoparticles for mouse ovary and cultured human granulosa cells. Ecotoxicology and Environmental Safety 249, 114371. 10.1016/j.ecoenv.2022.114371

Huang, Z., Weng, Y., Shen, Q., Zhao, Y., Jin, Y., 2021. Microplastic: A potential threat to human and animal health by interfering with the intestinal barrier function and changing the intestinal microenvironment. Science of The Total Environment 785, 147365. 10.1016/j.scitotenv.2021.147365

Jeong, B., Kim, J.-S., Kwon, A.R., Lee, J., Park, S., Koo, J., Lee, W.S., Baek, J.Y., Shin, W.-H., Lee, J.-S., Jeong, J., Kim, W.K., Jung, C.-R., Kim, N.-S., Cho, S.-H., Lee, D.Y., 2024. Maternal nanoplastic ingestion induces an increase in offspring body weight through altered lipid species and microbiota. Environment International 185, 108522. 10.1016/j.envint.2024.108522

Kannan, K., Vimalkumar, K., 2021. A Review of Human Exposure to Microplastics and Insights Into Microplastics as Obesogens. Frontiers in Endocrinology 12.

Kutralam-Muniasamy, G., Shruti, V.C., Pérez-Guevara, F., Roy, P.D., 2023. Microplastic diagnostics in humans: “The 3Ps” Progress, problems, and prospects. Science of The Total Environment 856, 159164. 10.1016/j.scitotenv.2022.159164

Leslie, H.A., van Velzen, M.J.M., Brandsma, S.H., Vethaak, A.D., Garcia-Vallejo, J.J., Lamoree, M.H., 2022. Discovery and quantification of plastic particle pollution in human blood. Environment International 163, 107199. 10.1016/j.envint.2022.107199

Lettieri, G., D’Agostino, G., Mele, E., Cardito, C., Esposito, R., Cimmino, A., Giarra, A., Trifuoggi, M., Raimondo, S., Notari, T., Febbraio, F., Montano, L., Piscopo, M., 2020. Discovery of the Involvement in DNA Oxidative Damage of Human Sperm Nuclear Basic Proteins of Healthy Young Men Living in Polluted Areas. International Journal of Molecular Sciences 21, 4198. 10.3390/ijms21124198

Levine, H., Jørgensen, N., Martino-Andrade, A., Mendiola, J., Weksler-Derri, D., Jolles, M., Pinotti, R., Swan, S.H., 2023. Temporal trends in sperm count: a systematic review and meta-regression analysis of samples collected globally in the 20th and 21st centuries. Human Reproduction Update 29, 157–176. 10.1093/humupd/dmac035

Lim, X., 2021. Microplastics are everywhere — but are they harmful? Nature 593, 22–25. 10.1038/d41586-021-01143-3

Liu, S., Shen, M., Li, C., Wei, Y., Meng, X., Li, R., Cao, Y., Wu, W., Liu, H., 2019. PKCδ contributes to oxidative stress-induced apoptosis in porcine ovarian granulosa cells via activating JNK. Theriogenology 131, 89–95. 10.1016/j.theriogenology.2019.03.023

Liu, Y., Ben, Y., Che, R., Peng, C., Li, J., Wang, F., 2023. Uptake, transport and accumulation of micro- and nano-plastics in terrestrial plants and health risk associated with their transfer to food chain - A mini review. Science of The Total Environment 902, 166045. 10.1016/j.scitotenv.2023.166045

Montano, L., 2020. Reproductive Biomarkers as Early Indicators for Assessing Environmental Health Risk, in: Hazardous Waste Management and Health Risks. Bentham Science Publisher, Sharjah, United Arab Emirates, pp. 113–145 (33).

Montano, L., Bergamo, P., Lorenzetti, M.G.A. and S., Montano, L., Bergamo, P., Lorenzetti, M.G.A. and S., 2018. The Role of Human Semen as an Early and Reliable Tool of Environmental Impact Assessment on Human Health, in: Spermatozoa - Facts and Perspectives. IntechOpen. 10.5772/intechopen.73231

Montano, L., Giorgini, E., Notarstefano, V., Notari, T., Ricciardi, M., Piscopo, M., Motta, O., 2023. Raman Microspectroscopy evidence of microplastics in human semen. Science of The Total Environment 901, 165922. 10.1016/j.scitotenv.2023.165922

Najahi, H., Alessio, N., Squillaro, T., Conti, G.O., Ferrante, M., Di Bernardo, G., Galderisi, U., Messaoudi, I., Minucci, S., Banni, M., 2022. Environmental microplastics (EMPs) exposure alter the differentiation potential of mesenchymal stromal cells. Environmental Research 214, 114088. 10.1016/j.envres.2022.114088

Oliveri Conti, G., Ferrante, M., Banni, M., Favara, C., Nicolosi, I., Cristaldi, A., Fiore, M., Zuccarello, P., 2020. Micro- and nano-plastics in edible fruit and vegetables. The first diet risks assessment for the general population. Environmental Research 187, 109677. 10.1016/j.envres.2020.109677

Pan, J., Liu, P., Yu, X., Zhang, Z., Liu, J., 2024. The adverse role of endocrine disrupting chemicals in the reproductive system. Front. Endocrinol. 14. 10.3389/fendo.2023.1324993

Park, Eun-Jung, Han, J.-S., Park, Eun-Jun, Seong, E., Lee, G.-H., Kim, D.-W., Son, H.-Y., Han, H.-Y., Lee, B.-S., 2020. Repeated-oral dose toxicity of polyethylene microplastics and the possible implications on reproduction and development of the next generation. Toxicol Lett 324, 75–85. 10.1016/j.toxlet.2020.01.008

Petro, E.M.L., Leroy, J.L.M.R., Covaci, A., Fransen, E., De Neubourg, D., Dirtu, A.C., De Pauw, I., Bols, P.E.J., 2012. Endocrine-disrupting chemicals in human follicular fluid impair in vitro oocyte developmental competence. Human Reproduction 27, 1025–1033. 10.1093/humrep/der448

Pironti, C., Notarstefano, V., Ricciardi, M., Motta, O., Giorgini, E., Montano, L., 2023. First Evidence of Microplastics in Human Urine, a Preliminary Study of Intake in the Human Body. Toxics 11, 40. 10.3390/toxics11010040

Pironti, C., Ricciardi, M., Motta, O., Miele, Y., Proto, A., Montano, L., 2021. Microplastics in the Environment: Intake through the Food Web, Human Exposure and Toxicological Effects. Toxics 9, 224. 10.3390/toxics9090224

Pulvirenti, E., Ferrante, M., Barbera, N., Favara, C., Aquilia, E., Palella, M., Cristaldi, A., Conti, G.O., Fiore, M., 2022. Effects of Nano and Microplastics on the Inflammatory Process: In Vitro and In Vivo Studies Systematic Review. FBL 27, 287. 10.31083/j.fbl2710287

Qian, N., Gao, X., Lang, X., Deng, H., Bratu, T.M., Chen, Q., Stapleton, P., Yan, B., Min, W., 2024. Rapid single-particle chemical imaging of nanoplastics by SRS microscopy. Proceedings of the National Academy of Sciences 121, e2300582121. 10.1073/pnas.2300582121

Rangel-Buitrago, N., Neal, W.J., 2023. A geological perspective of plastic pollution. Science of The Total Environment 893, 164867. 10.1016/j.scitotenv.2023.164867

Ricciardi, M., Pironti, C., Motta, O., Miele, Y., Proto, A., Montano, L., 2021. Microplastics in the Aquatic Environment: Occurrence, Persistence, Analysis, and Human Exposure. Water 13, 973. 10.3390/w13070973

Schell, T., Rico, A., Cherta, L., Nozal, L., Dafouz, R., Giacchini, R., Vighi, M., 2022. Influence of microplastics on the bioconcentration of organic contaminants in fish: Is the “Trojan horse” effect a matter of concern? Environmental Pollution 306, 119473. 10.1016/j.envpol.2022.119473

Shen, M., Lin, F., Zhang, J., Tang, Y., Chen, W.-K., Liu, H., 2012. Involvement of the Up-regulated FoxO1 Expression in Follicular Granulosa Cell Apoptosis Induced by Oxidative Stress*. Journal of Biological Chemistry 287, 25727–25740. 10.1074/jbc.M112.349902

Ullah, Sana, Ahmad, S., Guo, X., Ullah, Saleem, Ullah, Sana, Nabi, G., Wanghe, K., 2023. A review of the endocrine disrupting effects of micro and nano plastic and their associated chemicals in mammals. Frontiers in Endocrinology 13.

UNEP, 2021. United Nations Environment Programme (2021). Drowning in Plastics – Marine Litter and Plastic WasteVital Graphics.

Wang, D., Wang, W., Liang, Q., He, X., Xia, Y., Shen, S., Wang, H., Gao, Q., Wang, Y., 2018. DHEA-induced ovarian hyperfibrosis is mediated by TGF-β signaling pathway. Journal of Ovarian Research 11, 6. 10.1186/s13048-017-0375-7

Wang, J., Li, Y., Lu, L., Zheng, M., Zhang, X., Tian, H., Wang, W., Ru, S., 2019. Polystyrene microplastics cause tissue damages, sex-specific reproductive disruption and transgenerational effects in marine medaka (Oryzias melastigma). Environ Pollut 254, 113024. 10.1016/j.envpol.2019.113024

Wang, S., Han, Q., Wei, Z., Wang, Y., Xie, J., Chen, M., 2022. Polystyrene microplastics affect learning and memory in mice by inducing oxidative stress and decreasing the level of acetylcholine. Food and Chemical Toxicology 162, 112904. 10.1016/j.fct.2022.112904

Wei, Z., Wang, Y., Wang, S., Xie, J., Han, Q., Chen, M., 2022. Comparing the effects of polystyrene microplastics exposure on reproduction and fertility in male and female mice. Toxicology 465, 153059. 10.1016/j.tox.2021.153059

WHO, 2023. Infertility Prevalence Estimates, 1990–2021 [WWW Document]. URL https://www.who.int/publications-detail-redirect/978920068315 (accessed 3.28.24).

Wu, H., Liu, Q., Yang, N., Xu, S., 2023. Polystyrene-microplastics and DEHP co-exposure induced DNA damage, cell cycle arrest and necroptosis of ovarian granulosa cells in mice by promoting ROS production. Science of The Total Environment 871, 161962. 10.1016/j.scitotenv.2023.161962

Xu, W., Yuan, Yangyang, Tian, Y., Cheng, C., Chen, Y., Zeng, L., Yuan, Yuan, Li, D., Zheng, L., Luo, T., 2023. Oral exposure to polystyrene nanoplastics reduced male fertility and even caused male infertility by inducing testicular and sperm toxicities in mice. Journal of Hazardous Materials 454, 131470. 10.1016/j.jhazmat.2023.131470

Xue, T., Guan, T., Geng, G., Zhang, Q., Zhao, Y., Zhu, T., 2021. Estimation of pregnancy losses attributable to exposure to ambient fine particles in south Asia: an epidemiological case-control study. The Lancet Planetary Health 5, e15–e24. 10.1016/S2542-5196(20)30268-0

Yang, 2023. SciELO - Brazil - The impact of microplastics on female reproduction and early life The impact of microplastics on female reproduction and early life. Anim. Reprod., THEMATIC SECTION: 39TH ANNUAL MEETING OF THE ASSOCIATION OF EMBRYO TECHNOLOGY IN EUROPE (AETE) 20. 10.1590/1984-3143-AR2023-0037

Yang, W., Jannatun, N., Zeng, Y., Liu, T., Zhang, G., Chen, C., Li, Y., 2022. Impacts of microplastics on immunity. Front. Toxicol. 4. 10.3389/ftox.2022.956885

Yang, Z., Wang, M., Feng, Z., Wang, Z., Lv, M., Chang, J., Chen, L., Wang, C., 2023. Human Microplastics Exposure and Potential Health Risks to Target Organs by Different Routes: A Review. Curr Pollution Rep 9, 468–485. 10.1007/s40726-023-00273-8

Zhang, C., Chen, J., Ma, S., Sun, Z., Wang, Z., 2022. Microplastics May Be a Significant Cause of Male Infertility. Am J Mens Health 16, 15579883221096549. 10.1177/15579883221096549

Zhang, Y., Wang, X., Zhao, Y., Zhao, J., Yu, T., Yao, Y., Zhao, R., Yu, R., Liu, J., Su, J., 2023. Reproductive toxicity of microplastics in female mice and their offspring from induction of oxidative stress. Environmental Pollution 327, 121482. 10.1016/j.envpol.2023.121482

Zhao, J., Adiele, N., Gomes, D., Malovichko, M., Conklin, D.J., Ekuban, A., Luo, J., Gripshover, T., Watson, W.H., Banerjee, M., Smith, M.L., Rouchka, E.C., Xu, R., Zhang, X., Gondim, D.D., Cave, M.C., O’Toole, T.E., 2024. Obesogenic polystyrene microplastic exposures disrupt the gut-liver-adipose axis. Toxicological Sciences 198, 210–220. 10.1093/toxsci/kfae013

Zuccarello, P., Ferrante, M., Cristaldi, A., Copat, C., Grasso, A., Sangregorio, D., Fiore, M., Oliveri Conti, G., 2019a. Exposure to microplastics (<10 μm) associated to plastic bottles mineral water consumption: The first quantitative study. Water Research 157, 365–371. 10.1016/j.watres.2019.03.091

Zuccarello, P., Ferrante, M., Cristaldi, A., Copat, C., Grasso, A., Sangregorio, D., Fiore, M., Oliveri Conti, G., 2019b. Reply for comment on “Exposure to microplastics (<10 μm) associated to plastic bottles mineral water consumption: The first quantitative study by Zuccarello et al. [Water Research 157 (2019) 365–371].” Water Research 166, 115077. 10.1016/j.watres.2019.115077

Zurub, R.E., Cariaco, Y., Wade, M.G., Bainbridge, S.A., 2024. Microplastics exposure: implications for human fertility, pregnancy and child health. Front. Endocrinol. 14. 10.3389/fendo.2023.1330396

